# Effect of a theory-driven health education intervention on personal protective equipment use among commercial motorcycle riders in Cameroon: A quasi-experimental study

**DOI:** 10.64898/2026.04.08.26350441

**Authors:** Chrisantus Eweh Ukah, Nicholas Tendongfor, Alan Hubbard, Elvis Asangbeng Tanue, Rasheedat Oke, Nahyeni Bassah, Larissa Kumenyuy Yunika, Claudia Ngeha Ngu, S. Ariane Christie, Dickson Shey Nsagha, Alain Chichom-Mefire, Catherine Juillard

## Abstract

**Background:** Commercial motorcycle riders are among the most vulnerable road users in low- and middle-income countries and contribute substantially to the burden of road traffic injuries. The use of personal protective equipment (PPE), including helmets and protective clothing, reduces injury severity; however, uptake remains suboptimal. This study evaluated the effectiveness of a theory-driven health education intervention in improving knowledge, attitudes, and use of PPE among commercial motorcycle riders in Cameroon.

**Methods:** A quasi-experimental, non-randomized controlled before-and-after study was conducted in Limbe (intervention) and Tiko (control) Health Districts between August 4, 2024, and April 6, 2025. Participants were recruited from a cohort of commercial motorcycle riders and followed over an eight-month intervention period. The intervention, guided by the Health Belief Model and developed using the Intervention Mapping framework, combined face-to-face sensitization sessions with mobile phone–based educational messaging adapted to participants’ literacy levels and communication preferences. Data were collected at baseline and endline using structured questionnaires and direct observation checklists. Intervention effects were estimated using difference-in-differences analysis with generalized estimating equations, adjusting for socio-demographic factors.

**Results:** A total of 313 riders were enrolled at baseline (183 intervention, 130 control), with 249 retained at endline (149 intervention, 100 control). The intervention was associated with significant improvements in PPE knowledge (β = 2.91; 95% CI: 2.14–3.68; p < 0.001) and attitudes (β = 5.76; 95% CI: 4.32–7.21; p < 0.001) compared with the control group. No statistically significant effect was observed for PPE practice scores (β = 0.21; 95% CI: −0.09–0.52; p = 0.171). Among individual PPE items, helmet use increased significantly in the intervention group relative to the control group (AOR = 2.38; 95% CI: 1.19–9.45; p = 0.036), while no significant effects were observed for gloves, trousers, eyeglasses, or closed-toe shoes.

**Conclusion:** The theory-driven health education intervention significantly improved knowledge and attitudes toward PPE and increased helmet use among commercial motorcycle riders but did not lead to broader improvements in the uptake of other protective equipment. These findings highlight the need for complementary structural and policy interventions to address persistent barriers to PPE use in similar low-resource settings.

**Trial registration:** ClinicalTrials.gov Identifier: NCT07087444 (registered July 28, 2025, retrospectively)

## Background

Road traffic injuries (RTIs) remain a major public health challenge globally and are among the leading causes of death and disability, particularly among young and economically active populations[1,2]. The World Health Organization estimates that approximately 1.19 million people die each year due to road traffic crashes, with millions more sustaining non-fatal injuries, many resulting in long-term disability[3]. More than 90% of these deaths occur in low- and middle-income countries, with sub-Saharan Africa recording the highest regional mortality rates[4].

Motorcyclists represent one of the most vulnerable groups of road users and contribute substantially to the burden of RTIs[5]. In many low- and middle-income settings, motorcycles have become an essential mode of transport for both passengers and goods, particularly in urban and peri-urban areas where formal transport systems are limited. The rapid expansion of commercial motorcycle transport has been accompanied by an increase in motorcycle-related crashes and injuries, placing considerable strain on health systems and households[6,7].

Personal protective equipment (PPE), including helmets, protective jackets, gloves, trousers, and boots, plays a critical role in reducing the severity of injuries sustained during motorcycle crashes[8]. Evidence shows that helmet use alone can reduce the risk of death by approximately 40% and the risk of severe head injury by more than 70%[9]. Other protective clothing has also been associated with reductions in soft tissue injuries, fractures, and hospitalization among injured riders. Consequently, consistent and correct use of PPE is widely recognized as a cost-effective strategy for mitigating the health impact of motorcycle crashes[10].

Despite this evidence, the uptake of PPE among commercial motorcycle riders remains suboptimal in many low- and middle-income countries[8]. Previous studies across Africa and other low-resource settings have reported low levels of helmet use and minimal adoption of other protective gear. Factors such as limited knowledge, negative or indifferent attitudes, discomfort, cost, low availability, weak law enforcement, and social norms that normalize risky riding practices have been consistently identified as barriers to PPE use[11].

In Cameroon, road traffic injuries represent a growing public health concern, with motorcycles accounting for a large proportion of serious and fatal crashes, particularly in urban and peri-urban areas. Commercial motorcycle transport is widely used due to its affordability, flexibility, and ability to navigate poor road networks and traffic congestion. However, riders are frequently exposed to hazardous working conditions, including long riding hours, high traffic density, poor road infrastructure, and inconsistent enforcement of traffic regulations[12–14].

In the Limbe and Tiko Health Districts of the South-West Region of Cameroon, commercial motorcycle riding has become an important livelihood option, particularly in the context of rising unemployment and the ongoing socio-political crisis. Population displacement and economic instability have further increased reliance on motorcycle transport, leading to rapid growth in the number of riders and increasing reports of motorcycle-related injuries presenting to health facilities.

Although national regulations mandate helmet use in Cameroon, enforcement remains inconsistent, and the use of other forms of PPE is rarely promoted. Evidence from Cameroon and neighbouring countries suggests that many riders have limited knowledge of appropriate protective equipment, underestimate their risk of injury, and hold misconceptions regarding the effectiveness of PPE, contributing to poor compliance with safety practices[11].

Health education interventions have been recognized as an important component of comprehensive road safety strategies, particularly in low-resource settings where enforcement and structural interventions may be limited. However, the effectiveness of such interventions depends on their ability to address behavioural and contextual determinants of safety practices.

The Health Belief Model provides a useful framework for understanding PPE use among motorcycle riders by emphasizing perceived susceptibility to injury, perceived severity of outcomes, perceived benefits of protective actions, perceived barriers, cues to action, and self-efficacy[15]. In addition, the Intervention Mapping approach offers a systematic and participatory framework for designing context-specific interventions by integrating behavioural theory, empirical evidence, and stakeholder engagement[16,17].

Despite the importance of PPE and the potential of theory-based educational interventions, there is limited evidence from Cameroon evaluating the effectiveness of structured health education programs on PPE knowledge, attitudes, and use among commercial motorcycle riders. Most existing studies have focused on describing PPE use and associated factors, with few assessing whether targeted interventions can produce meaningful behavioural change.

This study therefore evaluated the effectiveness of a theory-driven health education intervention in improving knowledge, attitudes, and use of personal protective equipment among commercial motorcycle riders in the Limbe and Tiko Health Districts of Cameroon. The intervention was developed using the Health Belief Model and the Intervention Mapping framework and was designed to address locally identified barriers to PPE use. By generating empirical evidence in a crisis-affected setting, this study aims to inform scalable and context-appropriate injury prevention strategies for commercial motorcycle riders in Cameroon and similar settings.

## Materials and methods

### Study setting

The study was conducted in the Limbe and Tiko Health Districts, located in Fako Division of the South-West Region of Cameroon. The two districts are characterized by intense commercial motorcycle transport activity and serve as important economic and transport hubs in the region. Limbe is a coastal and touristic city, while Tiko hosts major agricultural and commercial enterprises and functions as a transit corridor between rural and urban communities. Both districts have experienced increasing population movement and economic instability related to the ongoing sociopolitical crisis in the North-West and South-West regions of Cameroon. As a result, commercial motorcycle riding has expanded rapidly as a livelihood option, increasing riders’ exposure to road traffic hazards.

### Specific Objectives and hypotheses

The primary objective of this study was to evaluate the effectiveness of a theory-driven health education intervention in improving the use of personal protective equipment (PPE) among commercial motorcycle riders in Limbe Health District compared with a control group in Tiko Health District. Specifically, the study aimed to assess the effect of the intervention on PPE-related knowledge, attitudes, and overall practices among riders, as well as to determine its effect on the use of individual PPE components, including helmets, gloves, trousers, eyeglasses, and closed-toe shoes.

It was hypothesized that the health education intervention would lead to a statistically significant increase in PPE use among commercial motorcycle riders in the intervention group compared with the control group over time. In addition, the intervention was expected to result in significant improvements in PPE-related knowledge and attitudes, as well as overall PPE practice scores. Furthermore, it was hypothesized that the intervention would lead to increased use of individual PPE components, particularly helmets, with smaller or variable effects anticipated for other PPE items such as gloves, trousers, eyeglasses, and closed-toe shoes.

### Study design

A quasi-experimental, non-randomized controlled before-and-after study design was used to evaluate the effect of a health education intervention on knowledge, attitudes, and use of personal protective equipment (PPE) among commercial motorcycle riders. Limbe Health District served as the intervention site, while Tiko Health District served as the control site. Allocation of districts to intervention and control arms was based on administrative and operational feasibility rather than randomization.

The same cohort of riders in both districts was followed from baseline to endline. The effect of the intervention was estimated using a difference-in-differences analytical framework to account for baseline differences and temporal trends between the two groups.

Ethical approval for the study was obtained prior to the commencement of data collection. The study protocol, including objectives, outcome measures, and analysis plan, was defined a priori before implementation of the intervention.

The study was registered with ClinicalTrials.gov (Identifier: NCT07087444; registered on July 28, 2025). The registration was completed retrospectively after the conclusion of data collection due to initial unawareness of trial registration requirements for quasi-experimental public health interventions. No changes were made to the study protocol, outcome measures, or analysis plan after the completion of the study.

### Study population

The study population consisted of commercial motorcycle riders operating within the Limbe and Tiko Health Districts. Riders were eligible for inclusion if they were aged 18 years or older, had been actively operating as commercial motorcycle riders within the study districts for at least three months, owned or operated a functional motorcycle, possessed a mobile phone, were able to communicate in English, French, or Pidgin English, and provided written informed consent. Riders who declined consent were excluded.

### Sampling and recruitment

Two comparable health districts within the same administrative division were purposively selected to minimize contextual differences. Limbe Health District was assigned as the intervention site, while Tiko Health District served as the control site.

The sampling frame for the intervention study was derived from participants who took part in a prior cross-sectional formative study conducted in the same districts[8]. During the cross-sectional study, commercial motorcycle riders were informed about a planned health education intervention. Riders who expressed willingness to participate in the intervention were identified, and their contact information, including phone numbers, was recorded.

A total of 300 riders in Limbe Health District and 199 riders in Tiko Health District were recruited during the cross-sectional study, forming the sampling frame for the intervention phase.

For the intervention study, eligible riders from this sampling frame were contacted by phone and invited to participate. Recruitment at baseline was therefore conducted from this pre-identified cohort of riders who had previously consented to be contacted. Of these, 183 riders in the intervention district (Limbe) and 130 riders in the control district (Tiko) were successfully enrolled at baseline.

Participants were followed over the intervention period, and endline data were collected among those who remained available and consented to follow-up. At endline, 149 riders in the intervention district and 100 riders in the control district were successfully retained.

The same eligibility criteria were applied at baseline and endline in both districts. Only riders who participated at baseline and consented to follow-up were included in the endline assessment. This approach minimized selection bias by ensuring that both intervention and control participants were drawn from the same underlying population.

### Flow of Participants Through the Study

**Figure 1:**
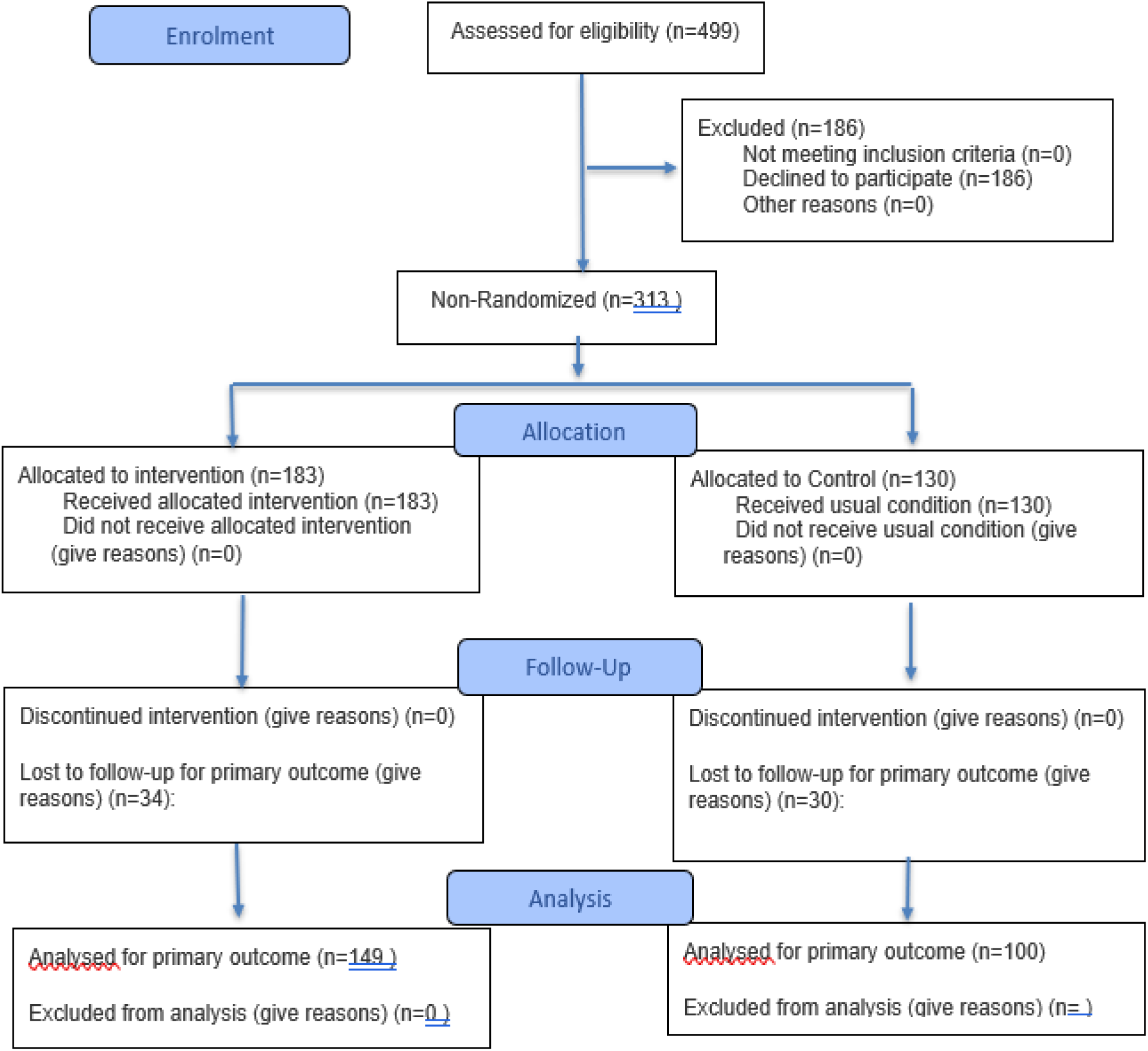
CONSORT flow diagram of participant recruitment, allocation, follow-up and analysis. Participants were identified from a prior cross-sectional study and subsequently recruited into the quasi-experimental intervention study based on willingness to participate. Allocation to study groups was non-randomized and based on study district. Citation: Hopewell S, Chan AW, Collins GS, Hróbjartsson A, Moher D, Schulz KF, et al. CONSORT 2025 Statement: updated guideline for reporting randomised trials. BMJ. 2025; 388:e081123. https://dx.doi.org/10.1136/bmj-2024-081123 © 2025 Hopewell et al. This is an Open Access article distributed under the terms of the Creative Commons Attribution License (https://creativecommons.org/licenses/by/4.0/), which permits unrestricted use, distribution, and reproduction in any medium, provided the original work is properly cited.

#### Sample size considerations

Sample size estimation was based on an anticipated improvement in PPE-related knowledge among commercial motorcycle riders following the health education intervention. A baseline knowledge proportion of 32.6%, a minimum detectable improvement of 16%, a 95% confidence level, and 90% statistical power were assumed. Allowing for loss to follow-up, a minimum of 100 riders per group was required. In practice, 183 riders were enrolled in the intervention group and 130 in the control group at baseline.

### Development of the health education intervention

The health education intervention was developed using a theory-driven and participatory approach informed by the Health Belief Model and the Intervention Mapping framework. The development process was grounded in findings from a formative qualitative needs assessment conducted among commercial motorcycle riders and key stakeholders in Limbe Health District.

### Formative needs assessment

A formative qualitative assessment was conducted prior to intervention development using focus group discussions and in-depth interviews with commercial motorcycle riders, motorcycle union leaders, road safety police officers, healthcare workers, and transport sector officials. This phase explored perceptions of road traffic injury risk, beliefs about PPE, barriers and facilitators to PPE use, and contextual factors influencing riding practices. Key barriers identified included low perceived susceptibility to injury, discomfort associated with PPE, cost and limited availability of protective equipment, misconceptions regarding PPE effectiveness, weak enforcement of road safety regulations, low literacy levels, and peer influence. Findings from this phase informed the development of intervention messages and delivery strategies.

### Theoretical framework

The intervention was guided by the Health Belief Model (HBM), a widely used behavioural theory for understanding and predicting health-related behaviours. The HBM posits that individuals are more likely to adopt protective behaviours when they perceive themselves to be susceptible to a health risk, believe the consequences of that risk are serious, perceive benefits in taking preventive action, and identify fewer barriers to performing the behaviour. In addition, cues to action and self-efficacy play critical roles in initiating and sustaining behavioural change.

In this study, the HBM informed the identification of key behavioural determinants influencing the use of personal protective equipment (PPE) among commercial motorcycle riders. Perceived susceptibility and severity were addressed by emphasizing riders’ risk of road traffic injuries and the potential consequences, including disability and death. Perceived benefits were highlighted through education on the protective effects of PPE in reducing injury severity, while perceived barriers such as discomfort, cost, and misconceptions about PPE effectiveness were explicitly addressed during intervention delivery. Cues to action were incorporated through repeated reminders via mobile phone messaging and periodic sensitization sessions, while self-efficacy was strengthened through demonstrations and practical guidance on proper PPE use.

To operationalize these theoretical constructs into a structured and context-specific intervention, the Intervention Mapping (IM) framework was applied. IM provided a systematic, stepwise approach to intervention development, including needs assessment, specification of behavioural and performance objectives, identification of theory-based methods, and translation of these methods into practical applications. Findings from the formative qualitative assessment were integrated into this process to ensure that the intervention addressed locally relevant barriers and facilitators of PPE use.

Through the combined application of the HBM and Intervention Mapping, the intervention was designed to not only increase knowledge but also influence attitudes and behavioural intentions, ultimately promoting sustained adoption of PPE among commercial motorcycle riders.

### Intervention content and delivery

The intervention was implemented exclusively in Limbe Health District over an eight-month period from August 4, 2024, to April 6, 2025. It was designed as a multi-component health education intervention combining face-to-face sensitization sessions with sustained mobile phone–based educational messaging to reinforce learning and promote behaviour change over time.

The face-to-face sensitization sessions were conducted by the Principal Investigator and project supervisors to ensure consistency, accuracy of information, and fidelity to the intervention design. Three group-based sensitization sessions were delivered at baseline, two months, and four months into the intervention period. These sessions were interactive in nature and included structured discussions on road traffic injury risks, the importance of personal protective equipment (PPE), and the consequences of non-use. Practical demonstrations were conducted to show correct use of PPE, including helmets and other protective gear. Participants were also given the opportunity to ask questions and clarify misconceptions, particularly those identified during the formative phase of the study, such as beliefs about discomfort, cost, and effectiveness of PPE.

To facilitate participation and improve attendance at the face-to-face sensitization sessions, minimal logistical support, including transportation reimbursement, was provided to participants. This was intended to reduce barriers to participation and ensure consistent exposure to the intervention.

To complement the face-to-face sessions and provide continuous reinforcement, mobile phone–based educational messages were delivered weekly throughout the intervention period. These messages were administered by trained research assistants who had at least a bachelor’s degree in a health-related field and were oriented on the content and objectives of the intervention. Messages were tailored to participants’ literacy levels and communication preferences and were delivered through multiple platforms, including SMS, WhatsApp text messages, and WhatsApp voice messages. The use of voice messages was particularly important for participants with low literacy levels.

The content of the messages focused on reinforcing key behavioural constructs, including increasing perceived risk of road traffic injuries, highlighting the benefits of PPE use, addressing common barriers, and providing cues to action. The repeated and multimodal delivery of messages was intended to enhance retention of information, sustain engagement, and support gradual adoption of safer riding practices.

### Control group

Commercial motorcycle riders in Tiko Health District served as the control group. No specific PPE-focused health education intervention was delivered in the control district during the study period. Participants continued to receive routine road safety information available in the district.

### Baseline and Endline Data Collection

Baseline data were collected in both districts prior to implementation of the intervention, and endline data were collected immediately after completion of the intervention. The same structured and pre-tested questionnaire was used during both baseline and endline surveys.

The questionnaire was developed specifically for this study based on relevant literature and findings from the formative qualitative phase and was pre-tested and refined prior to use.

The questionnaire collected information on socio-demographic characteristics, knowledge of personal protective equipment (PPE), attitudes toward PPE use, and self-reported use of PPE among commercial motorcycle riders. The full questionnaire is provided as Additional file 1.

In addition to self-reported data, direct observations were conducted at motorcycle parks and drop-off points using a structured checklist to assess the actual use of PPE during routine motorcycle operation. Riders were observed immediately after dropping off passengers to ensure that recorded behaviours reflected real-time PPE use rather than ownership or self-report.

The observational checklist captured the use of key PPE items, including helmets, gloves, trousers, protective eyewear, and closed-toe shoes. Data were collected electronically using Kobo Collect by trained data collectors. The same data collectors were used to collect data at baseline and endline to ensure consistency in measurement.

Interviews were conducted in English, with Pidgin English used where necessary to ensure comprehension and accuracy of responses.

### Measurement tools

Data were collected using two structured instruments developed specifically for this study: an interviewer-administered questionnaire and a direct observational checklist (Additional file 1).

The structured questionnaire was designed to collect information on participants’ socio-demographic characteristics, knowledge, attitudes, and self-reported practices related to personal protective equipment (PPE). The knowledge section consisted of multiple-choice and dichotomous questions assessing awareness of PPE types and their protective benefits. Attitudes toward PPE use were measured using Likert-scale items reflecting perceptions of risk, benefits, barriers, and willingness to use PPE. Practice-related questions assessed self-reported use of PPE during routine motorcycle riding.

The observational checklist was used to objectively assess actual use of PPE among riders. Observations were conducted immediately after riders dropped off passengers at designated points to ensure that recorded behaviours reflected real-time riding practices rather than ownership or self-report. The checklist captured the use of key PPE items, including helmets, gloves, trousers, protective eyewear, and closed-toe shoes.

Both instruments were developed based on relevant literature and informed by findings from the formative qualitative phase of the study. The tools were pretested in a similar setting, and necessary modifications were made to improve clarity and contextual relevance. Reliability of the questionnaire was assessed using Cronbach’s alpha, which yielded a coefficient of 0.78, indicating acceptable internal consistency.

### Outcome Measures

The primary outcome of this study was the change in the use of personal protective equipment (PPE) among commercial motorcycle riders. PPE use was assessed as both a composite practice score and as individual binary outcomes, including helmet use, gloves use, trousers use, eyeglasses use, and closed-toe shoes use, based on direct observation at baseline and endline.

Secondary outcomes included changes in knowledge and attitudes related to PPE use. Knowledge was assessed using a set of structured questions, with correct responses scored as one and incorrect responses as zero and summarized into a composite knowledge score.

Attitudes toward PPE use were measured using Likert-scale items and aggregated into a composite attitude score, with higher scores indicating more positive attitudes.

All outcomes were measured at baseline prior to the intervention and at endline following completion of the intervention. Continuous outcomes (knowledge, attitude, and practice scores) were analyzed as mean scores, while binary outcomes (individual PPE items) were analyzed as proportions. The effect of the intervention on these outcomes was assessed using a difference-in-differences analytical approach.

### Statistical analysis

All analyses were conducted using SPSS version 26. Descriptive statistics were used to summarize participant characteristics. Attrition analysis was performed to assess differences between participants retained and those lost to follow-up.

Within-group changes were assessed using paired samples t-tests for continuous outcomes and McNemar’s tests for binary outcomes. The intervention effect was estimated using a difference-in-differences approach implemented through generalized estimating equation models. Logistic GEE models were used for binary outcomes, and linear GEE models were used for continuous outcomes. Models included study group, time, and a group-by-time interaction term. Adjusted models controlled for age group, marital status, education level, and years of riding experience. Statistical significance was set at p < 0.05. Knowledge and attitude scores were treated as continuous variables.

### Post-intervention access for the control group

After completion of endline data collection, participants in the control district were provided access to the health education intervention for a period of two months using the same mobile phone–based delivery approaches.

### Ethical considerations

Ethical approval for this study was obtained from the Institutional Review Board of the Faculty of Health Sciences, University of Buea, Cameroon (Reference Number: 2024/2490-03/UB/SG/IRB/FHS). Administrative authorizations were also obtained from the Department of Public Health, University of Buea, the South-West Regional Delegation of Public Health, and the District Health Services of Limbe and Tiko prior to the commencement of the study. Participation was voluntary and written informed consent was obtained from all participants. For participants with limited literacy, the consent form was explained verbally in Pidgin English or a language understood by the participant. Confidentiality was maintained through the use of unique identifiers and secure databases. Participants were free to withdraw at any time. The study was conducted in accordance with the Declaration of Helsinki.

## Results

### Baseline socio-demographic characteristics of study Participants by study group

A total of 183 riders were enrolled in the intervention Health District (Limbe) at baseline and 149 were retained at endline. A total of 130 riders were enrolled at baseline in the controlled Health District (Tiko) and 100 were retained at the end of the intervention. A total of 249 commercial motorcycle riders were therefore retained at the end of the health education intervention study comprising 149 riders from Limbe Health District and 100 riders from Tiko Health District. The mean age of riders in the Limbe and Tiko Health Districts were 33.4±7.3 years and 32.0±7.8 years respectively. Most riders in both health districts were aged 31–40 years, accounting for 43.6% in Limbe and 39.0% in Tiko (Table 1). Overall, the age distribution of riders was broadly similar in both health districts, with the majority being younger adults.

**Table 1:** Socio-demographic characteristics of retained riders in the control and intervention groups.

| Variable | Category | Health District |  | Total |
| --- | --- | --- | --- | --- |
|  |  | Limbe | Tiko |  |
| Age group (years) | 41-50 | 25(16.8) | 8(8) | 33(13.3) |
|  | 31-40 | 65(43.6) | 39(39) | 104(41.8) |
|  | 21-30 | 58(38.9) | 50(50) | 108(43.4) |
| **Total** | **Total** | **149(99.3)** | **100(100)** | **249(100)** |
| Marital status | Married | 80(53.7) | 38(38) | 118(47.4) |
|  | Single | 69(45.3) | 62(62) | 126(50.6) |
| **Total** | **Total** | **149(100)** | **100(100)** | **249(100)** |
| Highest education | Tertiary | 14(9.4) | 5(5) | 19(7.6) |
|  | Secondary | 81(54.4) | 61(61) | 142(57) |
|  | Primary | 43(28.9) | 30(30) | 73(29.3) |
|  | No formal | 11(7.4) | 4(4) | 15(6) |
| **Total** | **Total** | **149(100)** | **100(100)** | **249(100)** |
| Average monthly income (XAF1000) | >150 | 11(7.4) | 8(8) | 19(7.6) |
|  | 101-150 | 47(31.5) | 27(27) | 74(29.7) |
|  | 50-100 | 85(57) | 61(61) | 146(58.6) |
|  | <50 | 6(4) | 4(4) | 10(4) |
| **Total** | **Total** | **149(100)** | **100(100)** | **249(100)** |
| Riding duration (years) | >10 | 26(17.4) | 17(17) | 43(17.3) |
|  | 5-10 | 48(32.2) | 29(29) | 77(30.9) |
|  | <5 | 75(50.3) | 54(54) | 129(51.8) |
|  | **Total** | **149(100)** | **100(100)** | **249(100)** |

### Assessment of loss to follow-up (attrition analysis) in the Control and Intervention Groups

An attrition analysis was conducted to assess differences between participants retained at endline and those lost to follow-up. The findings indicated that attrition was primarily associated with age groups, while other baseline characteristics were comparable between groups[18].

### Within-group comparisons between baseline and endline

#### 1. Within-group comparison of KAP scores in control and intervention groups

Paired samples t-tests were conducted to compare baseline and endline scores for knowledge, attitudes, and practices (KAP) related to personal protective equipment (PPE) among commercial motorcycle riders in both the control group (Tiko Health District) and the intervention group (Limbe Health District) (Table 2).

**Table 2:**
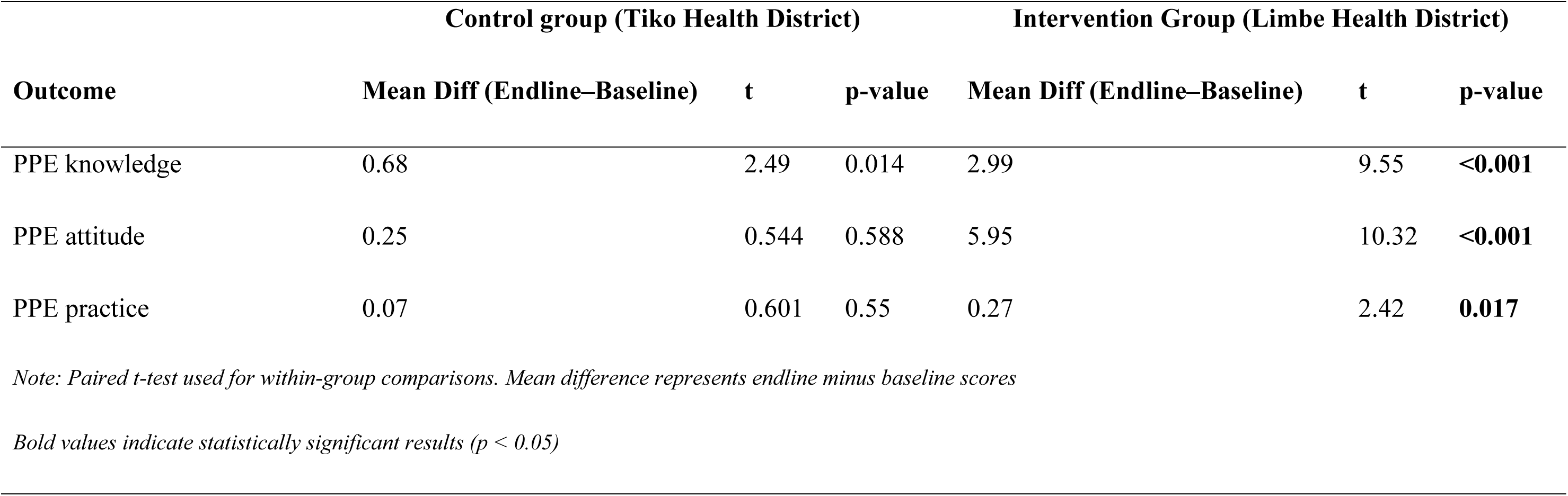
Within-group comparison of KAP scores in control and intervention groups.

Regarding the PPE outcomes in the control group, a statistically significant increase was observed only in knowledge scores (mean difference = 0.68, t = 2.49, p = 0.014). However, no statistically significant changes were found for PPE attitudes (mean difference = 0.25, t = 0.544, p = 0.588) or PPE practices (mean difference = 0.07, t = 0.601, p = 0.550).

In contrast, the intervention group demonstrated statistically significant improvements across all measured outcomes. There were statistically significant increases in knowledge (mean difference = 2.99, t = 9.55, p < 0.001), attitudes (mean difference = 5.95, t = 10.32, p < 0.001), and practices (mean difference = 0.27, t = 2.42, p = 0.017) toward PPE use. Overall, these findings indicate that while minimal changes were observed in the control group, the targeted health education intervention was associated with significant improvements in knowledge, attitudes, and practices related to PPE among riders in the intervention group.

#### 2. Within-group comparison of PPE use in control and intervention groups

McNemar’s test was used to compare changes in PPE use between baseline and endline among commercial motorcycle riders in both the control group (Tiko Health District) and the intervention group (Limbe Health District) (Table 3).

**Table 3:** Within-group comparison of PPE use in the control (Tiko) and the intervention (Limbe) groups.

| PPE Item | Control group (Tiko Health District, n=100) |  |  |  | Intervention group (Limbe Health District, n=149) |  |  |  |
| --- | --- | --- | --- | --- | --- | --- | --- | --- |
|  | Baseline (%) | Endline (%) | Change (%) | p-value | Baseline (%) | Endline (%) | Change (%) | p-value |
| Helmet | 20 | 23 | 3 | 0.736 | 22.8 | 34.2 | 11.4 | **<0.001** |
| Gloves | 18 | 18 | 0 | 1.000 | 16.8 | 18.8 | 2 | 0.766 |
| Trousers | 72 | 86 | 14 | **<0.001** | 83.9 | 89.9 | 6 | 0.200 |
| Eyeglasses | 8 | 9 | 1 | 1.000 | 9.4 | 19.5 | 10.1 | 0.024 |
| Closed-toe shoes | 81 | 84 | 3 | 0.25 | 81.9 | 87.2 | 5.4 | 0.302 |

In the control group, there were no statistically significant changes in the use of helmet (20.0% vs 23.0%, p = 0.736), gloves (18.0% vs 18.0%, p = 1.000), eyeglasses (8.0% vs 9.0%, p = 1.000), or closed-toe shoes (81.0% vs 84.0%, p = 0.250) between baseline and endline. However, a statistically significant change was observed in trousers use (72.0% vs 86.0%, p < 0.001). Despite this increase in percentage, further inspection of paired responses indicated that more riders discontinued the use of trousers than those who adopted it over time, suggesting an inconsistent pattern of change. Overall, PPE utilization remained largely unchanged in the control group.

In contrast, the intervention group demonstrated improvements in selected PPE components. There was a statistically significant increase in helmet use (22.8% vs 34.2%, p < 0.001) and eyeglasses use (9.4% vs 19.5%, p = 0.024) between baseline and endline. However, no statistically significant changes were observed for gloves (16.8% vs 18.8%, p = 0.766), trousers (83.9% vs 89.9%, p = 0.200), or closed-toe shoes (81.9% vs 87.2%, p = 0.302).

Overall, these findings suggest that while PPE use remained largely stable in the control group, the targeted health education intervention was associated with significant improvements in specific PPE components particularly helmet and eyeglasses use among riders in the intervention group.

To estimate the causal effect of the health education intervention on the use of personal protective equipment and visibility materials, a difference-in-differences (DiD) analytical approach was employed. The DiD method compares changes in outcomes over time in the intervention group (Limbe Health District) with changes over the same period in a comparable control group (Tiko Health District). This approach allows the effect of the intervention to be isolated from background temporal changes and other external influences that may affect both groups simultaneously, such as seasonal variations, enforcement activities or general public awareness activities.

Generalized estimating equations (GEE) were used to fit the DiD models in order to appropriately account for the correlated nature of the repeated measurements obtained from the same riders at baseline and endline. GEE provides population-averaged estimates and yields valid standard errors in the presence of within-subject correlation, making it particularly suitable for longitudinal and repeated-measures data. In this study, the intervention effect was represented by the interaction term between study group and time (Group × Time).

### Unadjusted difference-in-differences analysis of PPE use

Unadjusted difference-in-differences analyses using generalized estimating equations were conducted to assess the effect of the health education intervention on the use of personal protective equipment. The intervention was associated with a statistically significant increase in helmet use (COR = 2.31, 95% CI: 1.05–5.05, p = 0.036) No statistically significant intervention effects were observed for trousers use, eyeglasses use, closed-toe shoes use (all p > 0.05).

### Adjusted difference-in-differences analysis of PPE use

After adjusting for age group, level of education, average monthly income and riding experience, the health education intervention was associated with a statistically significant increase in helmet use (AOR = 2.38, 95% CI: 1.19–9.45, p = 0.036). No statistically significant intervention effects were observed for gloves use, trousers use, eyeglasses use, closed-toe shoes use (all p > 0.05). The adjusted models indicate that the health education intervention significantly improved helmet use among commercial motorcycle riders, while no significant effects were observed for the other PPE assessed (Table 4).

**Table 4:**
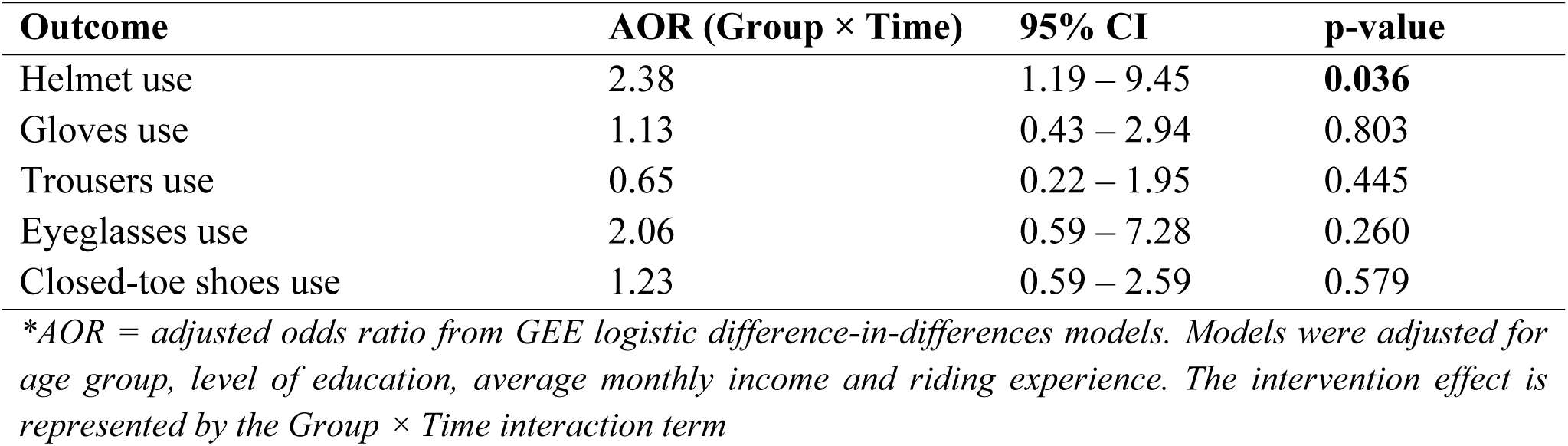
Adjusted difference-in-differences analysis of PPE use.

### Difference-in-Differences Analysis for Continuous Outcomes Unadjusted difference-in-differences analysis of KAP scores

Unadjusted difference-in-differences analyses using generalized estimating equations were conducted to assess the effect of the health education intervention on knowledge, attitudes and practices related to personal protective equipment. The intervention was associated with statistically significant increases in PPE knowledge scores (β = 2.91, 95% CI: 2.14–3.68, p < 0.001) and PPE attitude scores (β = 5.76, 95% CI: 4.32–7.21, p < 0.001). No statistically significant intervention effect was observed for PPE practice scores (β = 0.21, 95% CI: −0.09–0.52, p = 0.171).

### Adjusted difference-in-differences analysis of KAP scores

After adjusting for age group, marital status, level of education and years of riding, the health education intervention was associated with statistically significant improvements in knowledge and attitudes related to personal protective equipment (Table 5).

**Table 5:** Adjusted difference-in-differences (GEE) estimates of the effect of the intervention on KAP scores.

| Outcome | $\beta$ (Group × Time) | 95% CI | p-value |
| --- | --- | --- | --- |
| PPE knowledge score | 2.91 | 2.14 – 3.68 | <0.001 |
| PPE attitude score | 5.76 | 4.32 – 7.21 | <0.001 |
| PPE practice score | 0.21 | –0.09 – 0.52 | 0.171 |

*\*All models adjusted for age group, marital status, education and years of riding.*
*\* $\beta$ represents the difference-in-differences estimate from linear GEE models. The intervention effect is represented by the Group × Time interaction term.*

Specifically, PPE knowledge scores increased significantly in the intervention group compared with the control group (β = 2.91, 95% CI: 2.14–3.68, p < 0.001), as did PPE attitude scores (β = 5.76, 95% CI: 4.32–7.21, p < 0.001). However, no statistically significant intervention effect was observed for PPE practice scores (β = 0.21, 95% CI: −0.09–0.52, p = 0.171). These findings indicate that the health education intervention significantly improved riders’ knowledge and attitudes toward PPE but did not produce statistically significant changes in reported practices.

The similarity between the unadjusted and adjusted difference-in-differences estimates indicates that the measured socio-demographic variables did not confound the relationship between the health education intervention and the outcomes. The intervention effect therefore appears to be robust to adjustment for age, marital status, education and years of riding.

## Discussions

This study assessed the effectiveness of a theory-driven health education intervention on knowledge, attitudes, and use of personal protective equipment (PPE) among commercial motorcycle riders in Limbe and Tiko Health Districts. Guided by the Health Belief Model (HBM) and developed using the Intervention Mapping (IM) framework, the intervention resulted in significant improvements in riders’ knowledge and attitudes toward PPE and a statistically significant increase in helmet use. However, the intervention did not produce significant effects on the uptake of other PPE items, including gloves, trousers and closed-toe shoes.

The improvements observed in PPE-related knowledge and attitudes in the intervention group are consistent with the mechanisms proposed by the Health Belief Model[15,19]. The intervention explicitly targeted riders’ perceived susceptibility to road traffic injuries, perceived severity of injury outcomes, and perceived benefits of PPE use, while addressing commonly reported barriers such as discomfort, inconvenience and perceived low necessity of protective equipment. The statistically significant increases in PPE knowledge and attitude scores in the adjusted difference-in-differences models suggest that the intervention successfully modified riders’ cognitive and affective perceptions regarding PPE use.

The systematic use of the Intervention Mapping approach contributed to this outcome by ensuring that locally identified barriers and facilitators were directly translated into tailored educational messages and delivery strategies[17,20]. The formative qualitative phase revealed widespread misconceptions about the effectiveness of PPE, low perceived vulnerability to injury and limited trust in protective devices beyond helmets. These findings informed the design of culturally appropriate messages, demonstrations and repeated mobile phone reminders, which likely strengthened message relevance and acceptance among riders. This structured and participatory development process may explain the substantial gains in both PPE knowledge and attitudes observed in the intervention district.

Despite these improvements, the intervention effect on PPE practices was limited. Although within-group analyses showed significant improvements in PPE practice scores in the intervention group, the adjusted difference-in-differences analysis did not demonstrate a statistically significant overall intervention effect on PPE practices. This finding highlights an important and frequently observed gap between improved awareness and behavioural adoption in occupational safety and road safety interventions.

From an HBM perspective, this pattern suggests that although perceived susceptibility and perceived benefits may have increased, persistent perceived barriers continued to constrain behaviour change for several PPE items. During the formative phase, riders reported discomfort, heat, restricted movement and financial cost as major obstacles to consistent use of gloves, protective trousers and closed footwear. These barriers likely reduced riders’ perceived feasibility and self-efficacy for sustained use of multiple PPE items, even when positive attitudes were strengthened through the intervention.

Notably, helmet use emerged as the only PPE behaviour that improved significantly following the intervention in the adjusted models. Riders in the intervention district were more than twice as likely to use helmets after the intervention compared with the control group. This finding is particularly important in the study setting, where head injuries represent a major cause of morbidity and mortality among motorcyclists[9].

The stronger intervention effect observed for helmet use compared with other PPE items may be explained by several contextual and theoretical factors. First, helmets are widely recognized by riders as a core safety requirement and are more strongly embedded in existing traffic regulations and public messaging[21]. This normative environment likely reinforced the intervention’s cues to action and strengthened perceived benefits of helmet use. Second, helmets are often prioritized by riders because their protective role is well known, and their use is more visible to law enforcement, which may have increased perceived susceptibility to penalties for non-use[22]. These reinforcing environmental and regulatory cues are consistent with both the HBM and the environmental components emphasized in the Intervention Mapping framework.

In contrast, gloves, trousers and closed-toe shoes are not consistently enforced by traffic authorities and are often perceived primarily as comfort-related rather than safety-related equipment. The limited intervention effect on these items suggests that individual-level educational strategies alone may be insufficient to overcome entrenched behavioural and structural constraints[23]. This reinforces the Intervention Mapping principle that sustainable behaviour change requires alignment between individual determinants and environmental conditions[16].

The observed increase in eyeglasses use within the intervention group during the within-group analysis did not remain significant after adjustment in the difference-in-differences models. This suggests that part of the observed change may reflect temporal fluctuations or external influences affecting both districts rather than a direct intervention effect. This further highlights the importance of using controlled analytical approaches when evaluating behavioural interventions in dynamic real-world settings.

In the control district, a small but statistically significant improvement in PPE knowledge was observed over time, without corresponding changes in attitudes or practices. This may reflect general exposure to routine road safety messaging, informal peer discussions or media influences during the study period. However, the absence of meaningful behavioural change in the control group supports the conclusion that the observed increase in helmet use in the intervention district was attributable to the intervention rather than to background trends.

The findings of this study have important implications for PPE promotion among commercial motorcycle riders in similar low-resource and crisis-affected settings. While educational interventions grounded in behavioural theory can substantially improve knowledge and attitudes, translating these changes into consistent use of multiple PPE items may require complementary structural and policy-level interventions. Such measures could include improving access to affordable protective equipment, engaging motorcycle unions to establish collective safety norms, and strengthening enforcement of PPE regulations beyond helmet use[24].

Several limitations should be considered when interpreting these results. First, the quasi-experimental design cannot fully eliminate the possibility of unmeasured confounding. This was mitigated with the use of a difference-in-differences approach. Second, the intervention duration may not have been sufficient to support sustained adoption of more demanding or costly PPE behaviours, particularly in a setting characterized by economic hardship and unstable working conditions.

Nonetheless, this study provides evidence that a health education intervention developed through the Intervention Mapping framework and grounded in the Health Belief Model can significantly improve PPE-related knowledge, attitudes and helmet use among commercial motorcycle riders. Future interventions should incorporate multilevel strategies that address both individual beliefs and the structural barriers that constrain PPE uptake in order to achieve broader and more sustained improvements in rider safety.

## Conclusion

This study demonstrates that a theory-driven health education intervention, developed using the Health Belief Model and the Intervention Mapping framework, significantly improved commercial motorcycle riders’ knowledge and attitudes toward personal protective equipment in Limbe Health District. The intervention also resulted in a significant increase in helmet use compared with the control district. However, no significant intervention effects were observed for the use of other PPE items, including gloves, trousers and closed-toe shoes. These findings suggest that while structured and context-specific educational interventions can effectively influence cognitive and motivational determinants of PPE use, additional strategies addressing persistent practical and economic barriers are required to achieve broader and sustained adoption of multiple protective equipment items among commercial motorcycle riders in similar low-resource settings.

## Declarations

### Consent for publication

Not applicable.

### Data availability statement

All relevant data are within the manuscript and its Supporting Information files.

### Related work statement

This study is part of a broader research project evaluating the impact of a health education intervention among commercial motorcycle riders. A related manuscript focusing on visibility materials is currently under review elsewhere and is available as a preprint on Research Square[18]. The present manuscript focuses specifically on personal protective equipment outcomes, with distinct analyses and research objectives.

### Supplementary Information

**Additional file 1:** Structured questionnaire used for data collection (English version).

**Additional file 2:** Study protocol for the quasi-experimental intervention

**Additional file 3:** Related manuscript file

### Authors’ Contribution

**Conceptualization**: Chrisantus Eweh Ukah, Nicholas Tendongfor, Alan Hubbard, Elvis A. Tanue, Rasheedat Oke, Nahyeni Bassah, Sandra I. McCoy, Larissa Kumenyuy Yunika, Claudia Ngeha Ngu, S. Ariane Christie, Dickson S. Nsagha, Alain Chichom-Mefire, Catherine Juillard

**Data Curation**: Chrisantus Eweh Ukah, Nicholas Tendongfor, Alan Hubbard, Elvis A. Tanue, Rasheedat Oke, Nahyeni Bassah, Sandra I. McCoy, Larissa Kumenyuy Yunika, Claudia Ngeha Ngu, S. Ariane Christie, Dickson S. Nsagha, Alain Chichom-Mefire, Catherine Juillard

**Formal Analysis:** Chrisantus Eweh Ukah, Nicholas Tendongfor, Alan Hubbard, Elvis A. Tanue, Larissa Kumenyuy Yunika

**Funding Acquisition:** Ariane Christie, Sandra I. McCoy, Alain Chichom-Mefire, Catherine Juillard

**Investigation**: Chrisantus Eweh Ukah, Nicholas Tendongfor, Alan Hubbard, Elvis A. Tanue, Rasheedat Oke, Nahyeni Bassah, Sandra I. McCoy, Larissa Kumenyuy Yunika, Claudia Ngeha Ngu S. Ariane Christie, Dickson S. Nsagha, Alain Chichom-Mefire, Catherine Juillard

**Methodology:** Chrisantus Eweh Ukah, Nicholas Tendongfor, Alan Hubbard, Elvis A. Tanue, Rasheedat Oke, Nahyeni Bassah, Sandra I. McCoy, Larissa Kumenyuy Yunika, Claudia Ngeha Ngu, S. Ariane Christie, Dickson S. Nsagha, Alain Chichom-Mefire, Catherine Juillard

**Supervision:** Nicholas Tendongfor, Alan Hubbard, Catherine Juillard

**Validation:** Nicholas Tendongfor, Alan Hubbard, Elvis A. Tanue, Rasheedat Oke, Nahyeni Bassah, Ariane Christie,Catherine Juillard

**Visualization**: Chrisantus Eweh Ukah, Nicholas Tendongfor, Alan Hubbard, Elvis A. Tanue, Larissa Kumenyuy Yunika

**Writing – original draft:** Chrisantus Eweh Ukah, Nicholas Tendongfor, Alan Hubbard, Elvis A. Tanue, Rasheedat Oke, Nahyeni Bassah, Larissa Kumenyuy Yunika, Catherine Juillard

**Writing – Review and Editing:** Chrisantus Eweh Ukah, Nicholas Tendongfor, Alan Hubbard, Elvis A. Tanue, Rasheedat Oke, Nahyeni Bassah, Sandra I. McCoy, Larissa Kumenyuy Yunika, Claudia Ngeha Ngu, S. Ariane Christie^3^, Dickson S. Nsagha, Alain Chichom-Mefire, Catherine Juillard.

**Guarantor:** Chrisantus Eweh Ukah

**Patient and Public Involvement Statement**

Patients and the public were not involved in the design, conduct, reporting, or dissemination of this research. However, key stakeholders such as motorcycle union leaders and local health officers were consulted during the planning and data collection phases.

